# Impact of health system strengthening on delivery strategies to improve child immunization coverage and inequalities in rural Madagascar

**DOI:** 10.1101/2021.07.19.21260640

**Authors:** Elinanbinina Raojaonarifara, Matthew H. Bonds, Ann C Miller, Felana Ihantamalala, Laura F Cordier, Benedicte Razafinjato, Feno H. Rafenoarimalala, Karen E. Finnegan, Rado J. L. Rakotonanahary, Giovanna Cowley, Baolova Ratsimbazafy, Florent Razafimamonjy, Marius Randriamanambintsoa, Estelle M. Raza-Fanomezanjanahary, Andriamihaja Randrianambinina, C. Jessica E. Metcalf, Benjamin Roche, Andres Garchitorena

## Abstract

To reach global immunization goals, national programs need to balance routine immunization at health facilities with vaccination campaigns and other outreach activities (e.g. vaccination weeks), which boost coverage at particular times and help reduce geographic inequalities. However, where routine immunization is weak, an overreliance on vaccination campaigns may lead to heterogeneous coverage. Here, we assessed the impact of a health system strengthening (HSS) intervention on the relative contribution of routine immunization and outreach activities to reach immunization goals in rural Madagascar.We obtained data from health centers in Ifanadiana district on the monthly number of recommended vaccines (BCG, measles, DTP and Polio) delivered to children, during 2014-2018. We also analyzed data from a district-representative cohort carried out every two years in over 1500 households in 2014-2018. We compared changes inside and outside the HSS catchment in the delivery of recommended vaccines, population-level vaccination coverage, geographic and economic inequalities in coverage, and timeliness of vaccination. The impact of HSS was quantified via mixed-effects logistic regressions. The HSS intervention was associated with a significant increase in immunization rates (Odds Ratio between 1.22 for measles and 1.49 for DTP), which diminished over time. Outreach activities were associated with a doubling in immunization rates, but their effect was smaller in the HSS catchment. Analysis of cohort data revealed that HSS was associated with higher vaccination coverage (Odds Ratio between 1.18 per year of HSS for measles and 1.43 for BCG), a reduction in economic inequality, and a higher proportion of timely vaccinations. Yet, the lower contribution of outreach activities in the HSS catchment was associated with persistent inequalities in geographic coverage, which prevented achieving international coverage targets. Investment in stronger primary care systems can improve vaccination coverage, reduce inequalities and improve the timeliness of vaccination via increases in routine immunizations.

**Key questions:** *What is already known?:*

- Reaching the minimum recommended vaccination coverage of 90% for childhood illnesses remains a substantial challenge for low and middle income countries (LMICs).
- Understanding how vaccine delivery strategies can be improved to achieve coverage targets in rural areas of LMICs is essential due to the fragility of health systems and associated health budgets.
- While evidence exists on the impact of outreach activities and other targeted interventions aimed at improving immunization coverage, it is unclear how strengthening local health systems can help improve key indicators of vaccination coverage, via its different impacts on routine and outreach immunizations.

*What are the new findings?:*

- A health systems strengthening (HSS) intervention in a rural district of Madagascar improved overall vaccination coverage, reduced economic inequalities in vaccination coverage, and increased the proportion of timely vaccinations via an increase in routine immunizations.
- The contribution of outreach activities was lower in the HSS catchment area than in the rest of the district, which was associated with a persistence of geographic inequalities in vaccination coverage.

*What do the new findings imply?:*

- Strengthening local health systems can help improve key indicators of vaccination coverage in rural, low resource settings, even when those interventions do not target specifically vaccine improvements themselves.
- Explicit efforts are still necessary in areas undergoing HSS to vaccinate children in remote areas so that immunization goals can be reached.

## Introduction

Vaccination is one of the most effective public health interventions to reduce the burden of infectious diseases, particularly among children[1], [2]. To increase vaccination coverage around the world, the Global Alliance for Vaccines and Immunisation (GAVI) was created in 2000 to mobilize funds and technical expertise for child vaccination in the poorest countries in the world [3], [4]. As a result, from 2000 to 2015, global vaccination coverage has increased from 72% to 86% [5]. As of 2018, 760 million children have been immunized and an estimated 13 million deaths have been prevented in GAVI-supported countries [6]. Future impacts of immunizations are estimated to be larger with the introduction of new vaccines (e.g. rotavirus, papillomavirus) and the expansion of coverage for existing vaccines [7], [8]. Based on the Global Immunization Vision and Strategy, the goal of the Global Vaccine Action Plan is to reach a national coverage of 90% for basic vaccines in all countries by 2020 [8] [9], with at least 80% coverage in every district [10]. Despite great progress, vaccination coverage remains low in many areas of the developing world due to many reasons [11]. For instance, while average coverage for third dose of the DTP vaccine increased from 60% to 81% in low and middle income countries (LMICs), it remained under 40% for the bottom ten performing countries [5]. Failure to achieve critical population-level thresholds for herd immunity has resulted in sustained transmission, periodic epidemics and has slowed-down progress towards the elimination of vaccine preventable diseases such as polio, measles and rubella [12]–[14].

National strategies for vaccination in most low and middle income countries typically involve routine immunization (RI) at primary health centers, complemented with additional outreach activities to increase coverage such as periodic vaccination weeks (VW) and supplementary immunization activities (SIAs) such as mass vaccination campaigns. Routine immunization, where a child is brought to a health facility to receive the recommended shots, usually free of charge, represents the most reliable way of vaccinating children at the right time in order to maximize immunity [15]. However, its reach is undermined by the fragility of health systems in LMICs and multiple barriers faced by local populations for accessing health care [16]. In particular, geographic distance to primary health centers is associated with important inequalities in vaccination coverage [17]. Vaccination campaigns, which involve the mobilization of health workers to administer vaccines where populations live during VW and SIAs, are a very effective way to cover large geographic areas over short periods of time and to reduce geographic inequalities in vaccination coverage [18], [19]. Consequently, significant funding has been mobilized towards increasing coverage via vaccination campaigns [19], [20], but the low frequency of these campaigns can result in heterogeneous coverage across age groups [17], [21], insufficient number of recommended doses per vaccine [22], and important delays in immunization relative to the recommended age of vaccination [23]–[26]. In addition, vaccination campaigns can have negative impacts on subsequent rates of immunization via RI [27], [28], which could exacerbate these issues. To address this, investments in vaccination campaigns could be accompanied with broader health system strengthening (HSS) efforts to increase the contribution of RI to overall immunization coverage[29] [28].

Madagascar is illustrative of the challenges and potential solutions to achieving global goals for immunization in LMICs. Since its launch in 1976, the national Expanded Program on Immunization (EPI) has contributed to a substantial uptake in immunization [30], [31] which seems to have been an important driver in improvements in life expectancy [32]. In addition to supporting RI activities, the EPI launched biannual VWs in 2006 (“mother and child weeks”, which generally take place in April and November) [33]–[35], and conducts occasional SIAs to further increase coverage and prevent disease outbreaks [13], [36]. As of 2018, vaccination coverage goals for Madagascar had not yet been achieved for any of the recommended vaccines [37]. Suboptimal vaccination coverage can lead to larger-than-usual outbreaks (known in epidemiology as “post-honeymoon” epidemics) [13]. For instance, insufficient coverage for measles vaccine (∼80% by 2017) [38] led to the largest known measles outbreak in Madagascar history in 2018-2019 [39], which accounted for one fourth of global cases [41] that year with nearly 225 000 cases registered [40]. Achieving vaccination coverage targets is particularly challenging in rural areas of the country, where the majority of the population lives, and where coverage is over 10% lower than in urban areas [37] for all recommended vaccines.

In 2014, the Ministry of Public Health (MoPH) partnered with the nongovernmental healthcare organization, PIVOT, to strengthen the rural health district of Ifanadiana, located in southeastern Madagascar, to improve local health conditions and serve as a model health system for the country [42]. Though the partnership does not include a particular focus on immunization (which is managed directly by the MoPH), it supports a large range of interventions at health centers and community health sites in approximately one third of the district, which has resulted in substantial increases in primary health care access and utilization [43]. Those programs include improved ‘readiness’ of health facilities (staffing, training, equipment, infrastructure, supply chain) and clinical programs that can directly influence adherence to vaccinations schedules, such as family planning, antenatal care, postnatal care, and deliveries at health facilities.

The goal of this study was to assess the impact of HSS on the relative contribution of routine immunization and vaccination campaigns over time, and the impact of these changes on key features of immunization at the population-level. In particular, we assessed changes between 2014 and 2018 in the HSS catchment and in the rest of the district for a) the delivery of recommended vaccines, b) population-level vaccination coverage, c) geographic and economic inequalities in coverage, and d) timeliness of vaccination. For this, we combined immunization data from all health centers in Ifanadiana District with information from a district-representative longitudinal cohort conducted every two years in nearly 1600 households in the district (∼8000 individuals).

## Methodology

### Study site

Ifanadiana is a rural district in the region of Vatovavy Fitovinany, located in southeastern Madagascar. It comprises about 200 000 people distributed in 13 communes, with 2 additional communes created during the study period. The district’s health system consists of one hospital (CHRD) and at least one health center (CSB2) per commune that provides primary health care. Six communes have additional health centers (CSB1) with more limited health services. The initial HSS catchment comprised 4 out of the 13 communes in the first three years (2014 to 2016). One additional commune was added in 2017, with plans to progressively cover the entire district over the following years [44]. The HSS intervention spans across all levels of care (hospital, health centers and community health) and combines horizontal support to health system readiness (e.g. infrastructure, staffing, equipment, social support to patients) with vertical support to clinical programs (e.g. malnutrition, emergency care, tuberculosis) and improved information systems [44], [45]. More details are available in the Appendix, Table S1[46]. Delivery of child immunization is similar to the rest of Madagascar, combining RI with biannual VWs and other outreach activities. Only one SIA took place during the study period in Ifanadiana, a measles mass vaccination campaign in October 2016.

### Data collection

#### Health system data collection

Data on monthly immunization rates from 2014 to 2018 was obtained from all 19 primary health centers in Ifanadiana district. Two health centers that were recently built and lacked consistent data across the study period were excluded. Data was obtained on all recommended vaccines in the Madagascar EPI, which included tuberculosis (BCG), measles, polio, and the combined vaccine for diphtheria, tetanus and pertussis (DTP). For polio and DTP, only the number of third doses administered was considered. Immunization information was derived from the health centers’ monthly reports to the district, which are aggregated from the health centers’ registers every month by MoPH staff. From these, the number of children immunized per month for each of these vaccines was obtained for each health center (CSB1 or CSB2), which included all children vaccinated through both routine services and outreach activities. The population of children aged 12-23 months was also obtained for each health center catchment from official MoPH records [47]. Data quality was monitored by joint PIVOT-MoH supervisions every three months. Information on the geographic extent and timing of the HSS intervention was obtained from the NGO’s internal records.

#### Cohort data collection

We obtained population-level information from the Ifanadiana Health Outcomes and Prosperity longitudinal Evaluation (IHOPE), a district-representative longitudinal cohort study initiated in Ifanadiana district in 2014 [48]. It consists of a series of surveys conducted in a sample of 1600 households every two years, with questionnaires modeled after the internationally-validated Demographic and Health Surveys (DHS) [49]. A two-stage sample stratified the district by the initial HSS and control catchments. Eighty clusters, half from each stratum, were selected at random from enumeration areas mapped during the 2009 census, and households were then mapped within each cluster. Twenty households were selected at random from each cluster. 1522 households were successfully interviewed in 2014 (95.1% acceptance rate), 1514 and 1512 households were revisited during the follow-up in 2016 (94.6% acceptance rate) and in 2018 (94.5 % acceptance rate) respectively. Data collection, survey coordination and training were conducted by the Madagascar National Institute of Statistics (INSTAT).

The study was reviewed and approved by the Madagascar National Ethics Committee and the Harvard Medical School Institutional Review Board. The survey included a household questionnaire and individual questionnaires for all men and women of reproductive age (15-59 years and 15-49 years, respectively). All eligible women and men who were in the households sampled (usual residents or visitors) were interviewed. Data collected through the questionnaires included general information about household composition (size, genders, ages); living conditions, education, and other indicators of socio-economic status; recent illness, care seeking for illness and preventive behaviors; women’s reproductive history and care seeking behavior for reproductive health; children’s health, development, preventive behaviors, and care seeking for illness; and child, adult and maternal mortality. For vaccination specifically, information about vaccination status of the children under 5 years was obtained from the individual interviews with their mothers. Vaccination status and history was assessed from the children’s vaccination cards when available, or from the mother’s report otherwise.

### Data analysis

#### Analysis of immunization rates at health centers

First, for each vaccine (BCG, Polio 3^rd^ dose, DTP 3^rd^ dose, and Measles), we estimated monthly per capita rates (age-specific) at each health center. We then modelled temporal changes in immunization rates over the study period, studying the contribution of health system factors, time-varying factors and the effect of programs and policies. For health system factors, we studied baseline differences between health centers in the initial HSS catchment and in the rest of the district, as well as between different types of health center (CSB1 and CSB2). For time-varying factors, we studied annual linear and seasonal changes in immunization rates in the district. Seasonal changes were studied using a sine function with a period of one year and the horizontal shift that best fitted the data. For the effect of programs and policies, we studied the effect of VWs and of the HSS intervention on immunization rates. For this, we built dummy variables coded as 1 for the CSBs and months where each program was in place (discrete for months with VWs, and constant from the moment the HSS started until the end of the study period). We also studied the interaction of the HSS intervention with linear annual change and VWs, in order to account for the additional effect of the HSS intervention over time, and for changes in the contribution of VWs to overall immunization rates due to the HSS intervention. We excluded from the analysis the measles immunizations delivered via SIAs in October 2016 because the target age was children up to 5 years of age, which differed from the population group used in the analyses (12-23 months).

Per capita immunization rates were modelled separately for each vaccine via binomial regressions in generalized linear mixed models, including a random intercept for each health centers. All other variables were included as fixed effects. Univariate analyses were first performed for each explanatory variable and those with p-value below 0.1 were retained for multivariate analysis. From this full model, a reduced model that included only variables reaching statistical significance (p-value < 0.05) was obtained via backwards selection. Effects are reported as adjusted Odds Ratios (OR).

#### Analysis of vaccination coverage in the longitudinal cohort

While an analysis of health center immunizations can provide some basic understanding about the impact of the HSS intervention on RI and outreach activities over time, it does not allow for obtaining accurate measures of vaccination coverage due to known inaccuracies in target population estimates. In addition, aggregated information reported by the health system does not allow us to evaluate changes in economic or geographic inequalities in vaccination coverage, or for the assessment of the timeliness of vaccination, all of which can be affected by the relative contribution of RI and outreach activities in the area. For this, we conducted a complementary analysis of vaccination coverage at the population-level using data from the IHOPE cohort.

Vaccination coverage was estimated for 2014, 2016 and 2018 from individual level data for children 12-23 months or 12-59 months (depending on the analysis, see below), as the proportion of the target group immunized at the time of the interview. Similar to our analysis of health center immunization rates, we studied separately each of the recommended vaccines, namely BCG, Polio 3^rd^ dose, DTP 3^rd^ dose and measles. We also estimated whether the child had received all of these recommended vaccines. For each child surveyed, vaccination status for each vaccine was coded 1 if the child was vaccinated based either on the vaccination cards, or on the mother’s report, and 0 otherwise. To assess the impact of economic and geographic inequalities in vaccination coverage, we estimated a household wealth score via principal components analysis of household assets following standard DHS methods [49], and we estimated the shortest path distance from the villages in each cohort cluster to the nearest health center using the Open Source Routing Machine engine. For this, we had previously mapped the entire district of Ifanadiana on OpenStreetMap, resulting in over 23,000 km of footpaths and 5,000 residential areas mapped [50]. Households were ordered based on their wealth score and distance to the nearest health center and were classified into five quantiles with 20% of observations in each category (Q5 = wealthiest or closest to the health center; Q1 = poorest or most remote). Vaccination coverage in children 12-23 months was estimated inside and outside of the HSS catchment at the beginning and at the end of the study period (2014-2018), disaggregated by wealth quantile and by distance quantile. Changes in inequalities were measured as the gap in vaccination coverage between the worst-off quantiles (Q1-Q2) and the best-off quantiles (Q3-Q5) over time.

We then modelled changes in vaccination coverage over the study period, studying baseline differences and annual changes in overall coverage and in economic and geographic inequalities for the HSS catchment and the rest of the district. For this, we performed a separate logistic regression mixed model for each vaccine, with the household cluster as random intercept. To study baseline differences between HSS catchments we included a dummy variable reflecting whether clusters where located in the initial HSS catchment. We included the natural logarithm of the wealth score to study differences in socio-economic groups, and distance to health center (in tens of kilometers) to study differences in geographic groups, both as continuous variables. We included two time-varying variables, one to reflect annual changes in vaccination coverage in the whole district, and another to reflect changes per year of HSS intervention in the HSS catchment. Finally, we included interaction terms of these two variables with wealth and distance to study the evolution of inequalities in each area. We included children aged 12-59 months in these analyses to allow for adequate sample sizes for each model. Model selection procedures were identical to those described above for the analysis of health system data. To understand which population groups could reach recommended vaccination coverage targets in the HSS catchment and in the rest of the district, we predicted in-sample vaccination coverage for 2018 from each of the reduced multivariate models, at varying levels of socio-economic class and proximity to health centers.

Finally, we studied the difference in timeliness of vaccination between the HSS catchment and the rest of the district in the subset of children 12-59 months with vaccination cards at the time of the interview in any of the cohort waves (N=786). For this, we estimated the child’s age at vaccination from the date of birth and the date of vaccination. Timely vaccination was estimated for each vaccine based on the recommended age of vaccination by the national EPI in Madagascar: in the first month of life for BCG (recommended to be given at birth), in the fourth month for Polio 3^rd^ dose and DTP 3^rd^ dose (recommended to be given the 14^th^ week), and in the ninth month for measles [37].

## Results

### Trends in the rates of per capita immunization at Ifanadiana’s health centers

Between January 2014 and December 2018, a total of 28,407 BCG, 31,476 Polio 3^rd^ dose, 33,241 DTP 3^rd^ dose, and 30,371 measles immunizations were delivered by the 19 health centers in Ifanadiana District. Average monthly per capita immunization rates (age specific, children 12-23 months) at health centers varied from 0.02 to 0.21, with an average of 0.08. Higher rates were observed on average in the HSS catchment, during months where VWs took place, and with an apparent increase over time in the whole district (Figure 1). These immunization trends were similar for all the different vaccines considered (Figure 1). Results from multivariate analyses revealed that per capita immunization rates were similar for different types of health center and HSS catchment at baseline (Table 1). Immunization rates for all vaccines increased over time and varied seasonally, with higher rates during the dry season (peak in August) and lower rates during the rainy season (bottom in February). Annual increase was higher for BCG and measles (OR 1.23 and 1.1 respectively), which require one single dose, than for polio and DTP (OR=1.06 for both), which require three doses. VWs were associated with approximately a doubling in immunization rates in the months where they took place (OR between 1.88 measles and 2 for polio).

**Table 1.** Determinants of per capita monthly immunizations at health centers in Ifanadiana district, 2014-2018 (GLMM, multivariate results^1^).

| Variable | BCG<br>immunizations | Polio<br>immunization<br>(3rd dose) | DTP<br>immunization<br>(3rd dose) | Measles<br>immunization |
| --- | --- | --- | --- | --- |
| Monthly coverage at baseline (intercept) | 0.04 (0.03-0.04) | 0.06 (0.06-0.07) | 0.06 (0.06-0.07) | 0.06 (0.05-0.06) |
| **Time-varying factors** |  |  |  |  |
| Annual change | 1.23 (1.22-1.25) | 1.06 (1.05-1.07) | 1.06 (1.05-1.07) | 1.1 (1.09-1.11) |
| Seasonal changes | 1.05 (1.03-1.07) | 1.05 (1.03-1.07) | 1.06 (1.04-1.08) | 0.98 (0.96-1) |
| **Effect of programs and policies** |  |  |  |  |
| Mother and child week (2 months per year) | 1.95 (1.89-2.02) | 2 (1.93-2.06) | 1.95 (1.89-2.01) | 1.88 (1.82-1.95) |
| Health system strengthening (HSS) | 1.4 (1.29-1.52) | 1.34 (1.25-1.44) | 1.49 (1.39-1.6) | 1.22 (1.14-1.32) |
| HSS x Annual change | 0.95 (0.93-0.97) | 0.92 (0.91-0.94) | 0.91 (0.9-0.93) | 0.92 (0.91-0.94) |
| HSS x Mother and child weeks | 0.73 (0.69-0.77) | 0.62 (0.58-0.66) | 0.65 (0.61-0.69) | 0.77 (0.73-0.82) |

<sup>1</sup> Results are expressed as probabilities for the intercept and as Odds Ratio with associated 95% confidence intervals for all other variables. Models initially controlled for health system factors (type of health center and baseline differences in HSS catchment vs. control) but these were removed in the final reduced models for lack of statistical association

**Figure 1.**
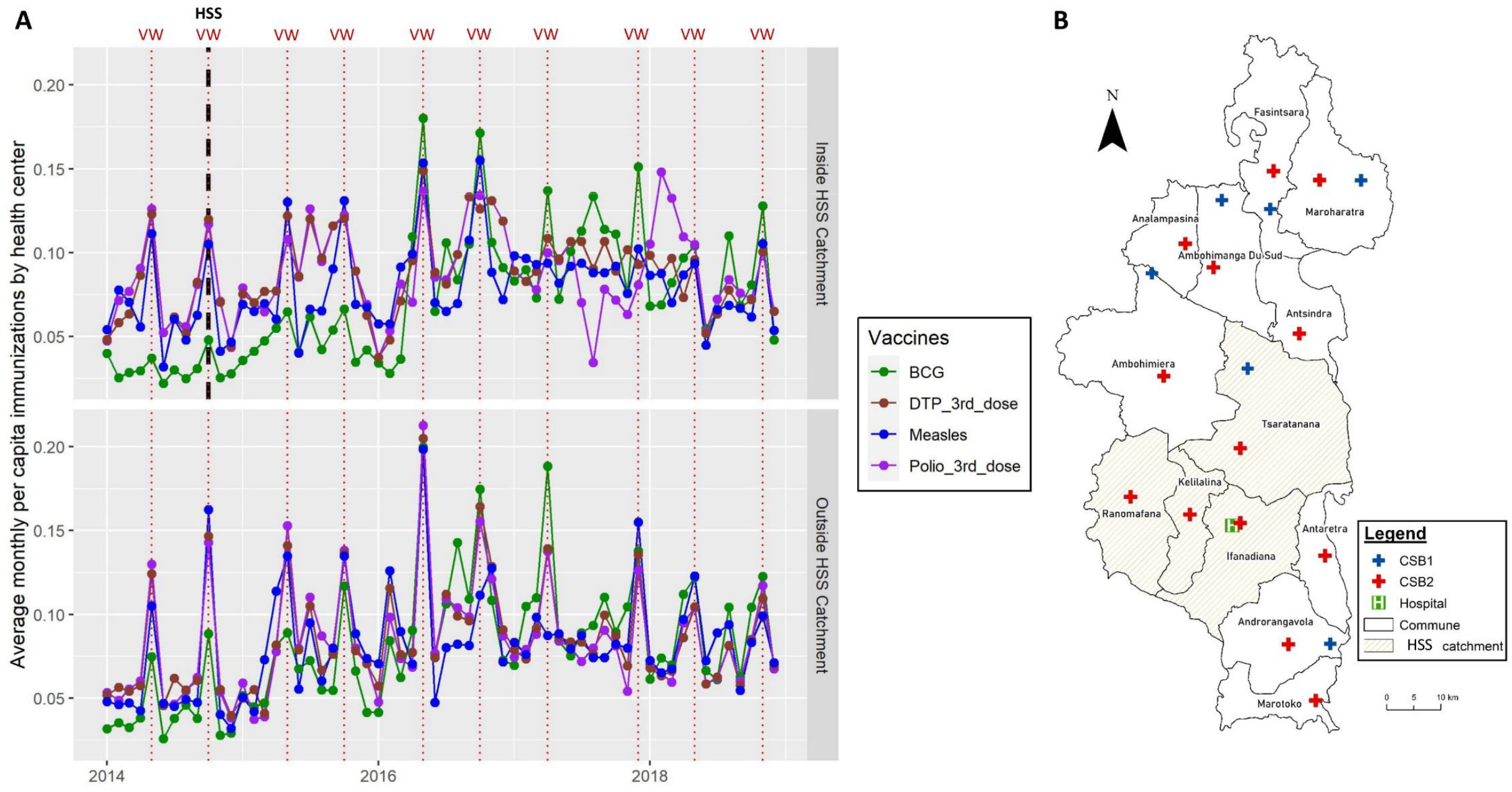
Changes in monthly immunization rates for children 12-23 months at health facilities in Ifanadiana District, 2014-2018. **A)** Average number of monthly immunizations per capita (age-specific, 12-23 months) delivered by health centers over time in the HSS catchment and in the rest of the district, with colors representing different vaccines. **B)** Map of Ifanadiana District and its health facilities. The initial HSS catchment is shown as yellow shaded areas, whereas the rest of the district is shown as white areas.

The HSS intervention, implemented since October 2014 in a third of Ifanadiana district, was associated with a significant increase in immunization rates (OR between 1.22 for measles and 1.49 for DTP), although this effect diminished over time (OR for interaction of HSS with annual change between 0.91 for DTP and 0.95 for BCG). Interestingly, the relative contribution of VWs to overall immunization rates was lower in the HSS catchment following the HSS intervention, with an OR for the interaction with VWs between 0.62 for polio and 0.77 for measles (Table 1).

### Changes in population-level vaccination coverage from the longitudinal cohort

#### Trends in vaccination coverage and inequalities

In total, data from 2,699 children between 12 and 59 months of age was obtained from the longitudinal cohort. Of these, 651 were between 12 and 23 months old, the age at which all four immunizations studied here should be completed. Vaccination coverage for children 12-23 months was very low at baseline, ranging from about 54 to 59% depending on the vaccine. Only 34.6% of children 12-23 months were fully vaccinated in 2014. Consistent with analyses of health system data, coverage for most vaccines improved substantially during the study period, especially in the HSS catchment (Figure 2). In 2018, 63.6% of children were fully vaccinated in the HSS area, compared with only 37.5% in the rest of the district. Coverage in 2018 varied for each vaccine considered; BCG had the highest coverage (80.8% inside and 70.3% outside the HSS catchment), whereas measles had the lowest coverage (73.2% inside and 49.2% outside the HSS catchment). The minimum recommended coverage of 90% was not reached for any of the vaccines, either inside or outside the HSS catchment.

**Figure 2.**
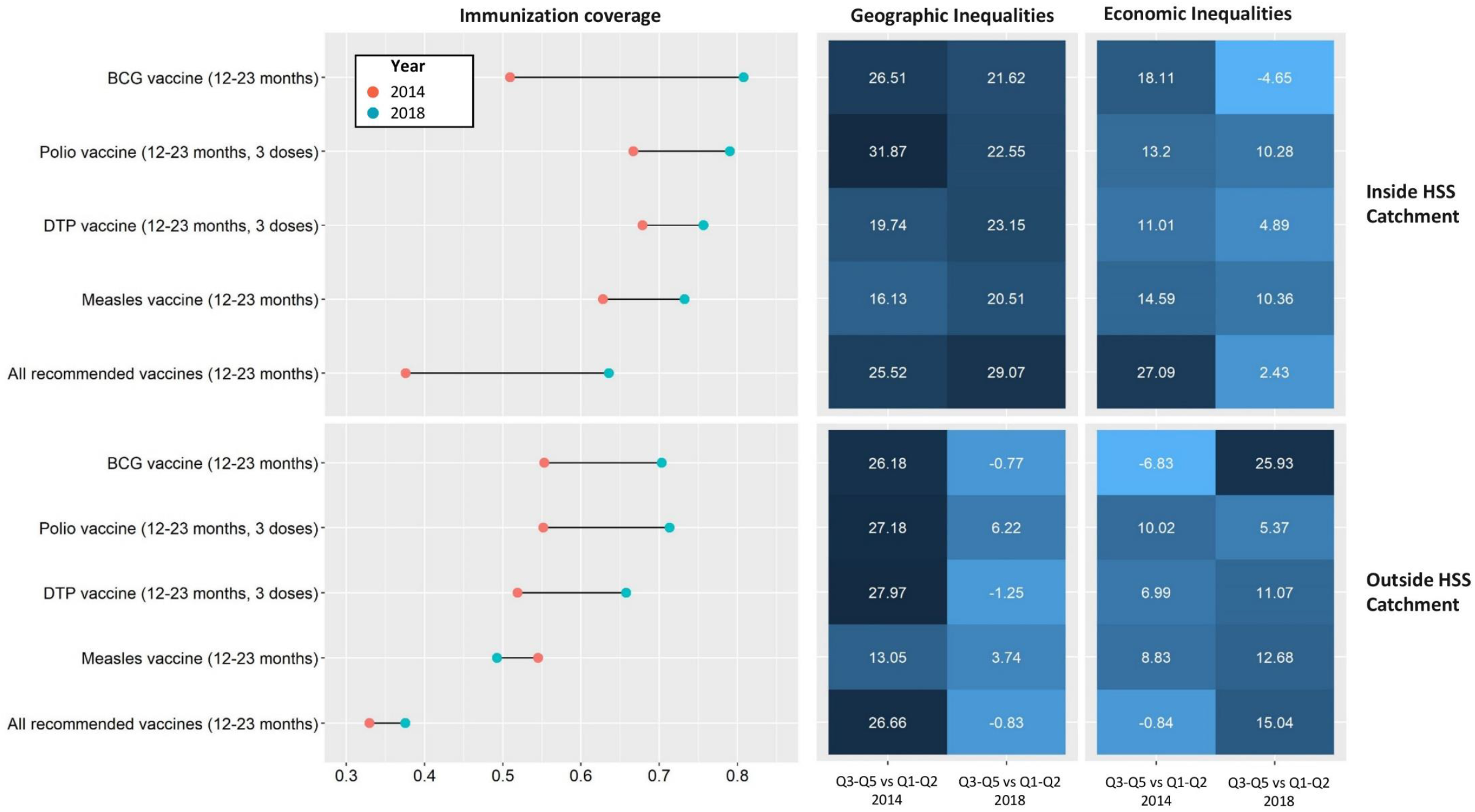
Changes in vaccination coverage for children 12-23 months and associated inequalities in Ifanadiana District, 2014-2018. Left panel shows changes in immunization coverage over time, split by HSS catchment and type of vaccine. Middle and right panels show inequalities in coverage over time, according to geographic quantiles (distance to health center) and economic quantiles (wealth score), respectively. Color gradient shows the difference in coverage between the best-off groups (quantiles 3 to 5) and the worst-off groups (quantiles 1 and 2), from dark blue (greater difference, more inequalities) to light blue (smaller difference, less inequalities).

Disparities in immunization coverage were observed according to households’ geographic distance to health centers and wealth, with different trends in the HSS catchment and in the rest of the district (Figure 2). In 2014, the difference in coverage between households living closer (quantiles Q3-Q5) and further (Q1-Q2) from health centers ranged from 25% to 32%, except for measles vaccine. Differences between wealthier (Q3-Q5) and poorer (Q1-Q2) households were smaller, between 5 and 15% for most vaccines. After four years, economic inequalities in vaccination coverage were substantially reduced in the HSS catchment, with little change in geographic inequalities. In contrast, in the rest of the district geographic inequalities were greatly reduced, while economic inequalities increased for all vaccines except for polio. Table S2 shows vaccination coverage rates in each of the cohort years (2014, 2016, and 2018) and different population groups.

#### Determinants of vaccination coverage trends and predictions of coverage targets

Multivariate analyses of vaccination coverage trends between 2014 and 2018 revealed consistent predictors for most of the vaccines studied (Table 2). Baseline differences between the HSS catchment and the rest of the district were observed for only two vaccines, BCG (OR=0.6) and DTP (OR=1.65). Coverage of each of the four vaccines was positively associated with household wealth and negatively associated with household distance to health centers. The odds of vaccination for children in remote households was between half (OR=0.52, measles) and a fifth (OR=0.22, BCG) for every additional 10 km from the nearest health center. Vaccines with three required doses were the most associated with household wealth, with an odds ratio of 2.58 for DTP and 2.85 for polio. District-wide improvements in vaccination coverage were associated with a reduction in geographic inequalities over time and not with a homogeneous improvement for all population groups. Indeed, the OR of the interaction between annual change and distance to health center ranged from 1.17 (all vaccines) and 1.31 (BCG and polio), meaning that each year households living far from health centers had progressively better coverage, closing the gap with those living in close proximity. Changes in the HSS catchment were distinct from the rest of the district. First, every year of HSS intervention was associated with an increase in the odds of vaccination in the HSS catchment between 1.18 (measles) and 1.43 (BCG), except for DTP. Unlike the rest of the district, children from wealthier households in the HSS catchment had lower odds of vaccination over time (OR of interaction ranging from 0.73 to 0.83). However, the decrease in the odds of vaccination over time for more remote households in the HSS catchment (OR of interaction ranging from 0.72 to 0.84) effectively compensated the positive effect observed in the district as a whole, meaning that geographic inequalities were only reduced outside the HSS catchment.

**Table 2.** Determinants of vaccination coverage at the population level in Ifanadiana district, 2014-2018 (GLMM, multivariate results^1^).

| Variable | BCG<br>immunization | Polio<br>immunization<br>(3rd dose) | DTP<br>immunization<br>(3rd dose) | Measles<br>immunization | All<br>recommended<br>vaccines |
| --- | --- | --- | --- | --- | --- |
| Immunization coverage at baseline (intercept) | 0.8 (0.73-0.86) | 0.77 (0.71-0.83) | 0.71 (0.62-0.78) | 0.68 (0.61-0.75) | 0.47 (0.38-0.56) |
| **District-wide differences** | - | - | - | - | - |
| Baseline differences in HSS catchment vs. control | 0.6 (0.39-0.92) | - | 1.65 (1.12-2.44) | - | - |
| Socio-economic class (log of wealth score) | 2.18 (1.51-3.15) | 2.85 (1.92-4.23) | 2.58 (1.75-3.8) | 2.3 (1.6-3.32) | 2.68 (1.84-3.91) |
| Distance to health center (every 10 km) | 0.22 (0.12-0.4) | 0.3 (0.17-0.51) | 0.31 (0.18-0.53) | 0.53 (0.33-0.85) | 0.35 (0.19-0.62) |
| **Changes in the district** |  |  |  |  |  |
| Annual change | - | - | - | - | - |
| Annual change x Socio-economic class | - | - | - | - | - |
| Annual change x Distance to health center | 1.31 (1.21-1.41) | 1.31 (1.21-1.41) | 1.23 (1.14-1.33) | - | 1.17 (1.08-1.27) |
| **Changes in the HSS catchment** |  |  |  |  |  |
| Change per year of HSS | 1.43 (1.22-1.66) | 1.19 (1.04-1.36) | - | 1.18 (1.04-1.34) | 1.22 (1.08-1.38) |
| Change per year of HSS x Socio-economic class | - | 0.75 (0.6-0.95) | 0.83 (0.67-1.03) | 0.78 (0.63-0.97) | 0.73 (0.59-0.9) |
| Change per year of HSS x Distance to health center | 0.81 (0.65-1) | 0.72 (0.58-0.9) | 0.84 (0.73-0.96) | 0.76 (0.62-0.91) | 0.81 (0.66-1) |

<sup>1</sup> Results are expressed as probabilities for the intercept and as Odds Ratio with associated 95% confidence intervals for all other variables. A sign “-” means that the variable was not part of the final reduced model for lack of statistical association.

In-sample predictions from these multivariate models for 2018 revealed stark differences for achieving international coverage targets depending on HSS support and population characteristics (Figure 3). Overall, a 90% coverage (recommended coverage at the national level) could only be achieved for BCG, and just for populations who live in close proximity to a health center with HSS support and who are among the wealthiest in the area. When the target is relaxed to 80% coverage (minimum coverage recommended for every district), there were some population subgroups for which this target could be achieved in the HSS catchment for every individual vaccine. The range of socio-economic and geographic groups for which minimum coverage rates could be reached was much larger for BCG and Polio than for DTP and measles (Figure 3). Coverage targets for all recommended vaccines simultaneously (instead of each independently) could not be achieved for any population group. In areas outside of the HSS catchment, a 90% coverage was not achieved for any of the recommended vaccines or population subgroups. Only those in the top percentiles of wealth and proximity to a health center achieved an 80% coverage for BCG vaccination without HSS support.

**Figure 3.**
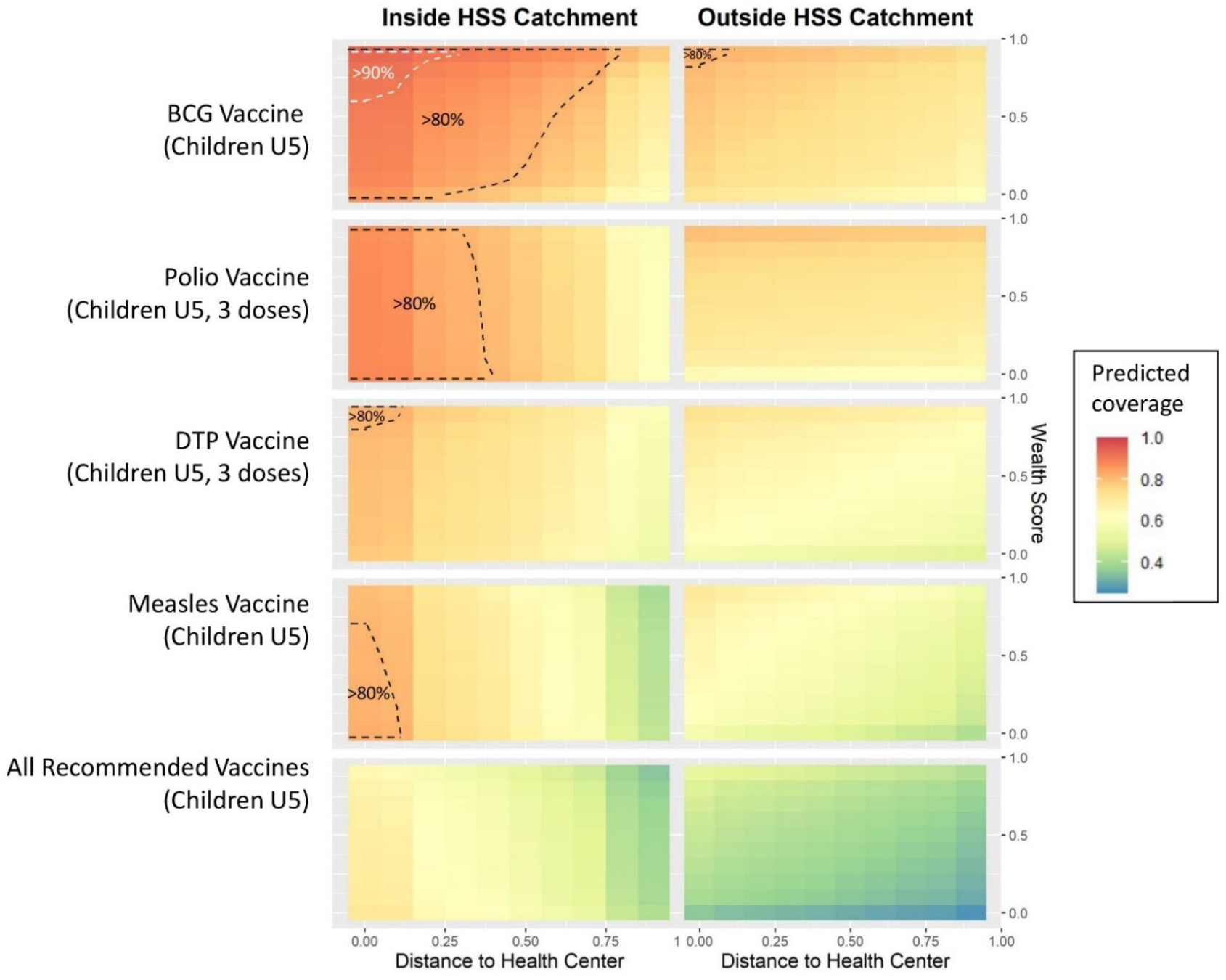
Predictions for achieving vaccination coverage targets for different population groups in Ifanadiana District. Graphs show in-sample predictions of vaccination coverage for the year 2018, estimated from models fitted with the cohort dataset (coefficients available in Table 2). For this, vaccination coverage was estimated for every combination of household distance to health center and wealth (split into deciles) in the HSS catchment and in the rest of the district, using the fixed effects of each model. Areas with predicted coverage greater than 90% or 80% are surrounded with white dashed lines or black dashed lines, respectively.

#### Timeliness of vaccination

Among the 786 children aged 12-59 months who had a vaccination card at the time of the interview (of children), timeliness of vaccination varied widely depending on HSS support and the vaccine considered (Figure 4). Most children were vaccinated in the first month of life for BCG, at 4-5 months for the third dose of polio and DTP, and at 9-10 months for measles (Figure 4A). Vaccination occurred later than recommended in national policies (see methods section) for many children, especially outside the HSS catchment. As a result, the proportion of children vaccinated at the recommended age was higher in the HSS catchment, ranging between 58% for BCG and 44% for polio and DTP (Figure 4B). In the rest of the district, this proportion was significantly lower and ranged between 49% for BCG and 22% for polio and DTP. Timeliness of vaccination improved between 2014 and 2018 in the HSS catchment for all vaccines except for BCG, while it only improved for measles in the rest of the district (Figure S1).

**Figure 4.**
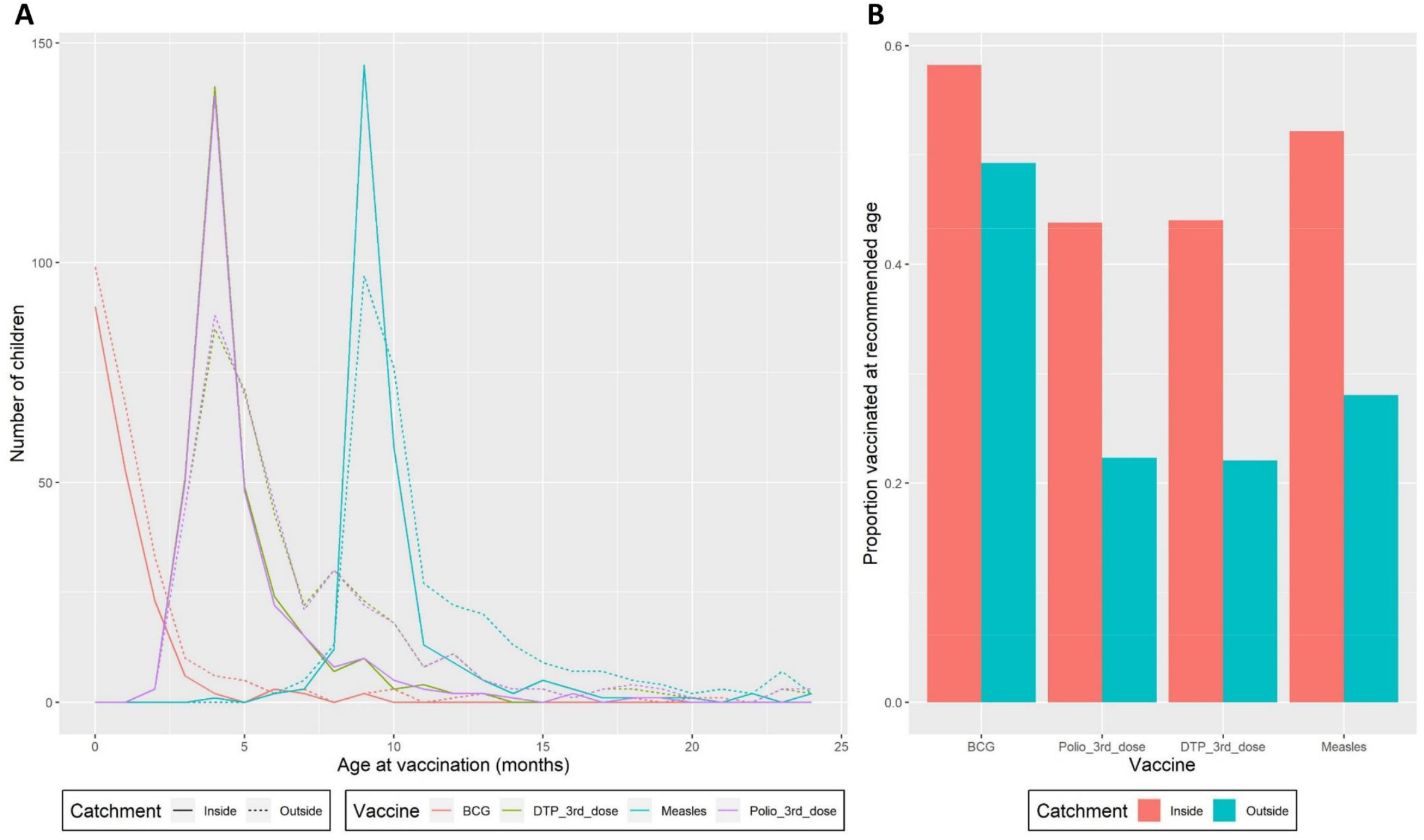
Timeliness of vaccination among children 12-59 months in Ifanadiana District. **A)** Age of vaccine administration (in months) as reported in children’s vaccination cards inside the HSS catchment (solid lines) and in the rest of the district (dashed lines), with colors representing each type of vaccine. B) Proportion of children vaccinated at the recommended age for BCG (1^st^ month), third dose of Polio (4^th^ month), third dose of DTP (4^th^ month) and Measles (9^th^ month), as per the Madagascar Expanded Program on Immunization.

## Discussion

The COVID-19 pandemic has brought renewed attention to the benefits and challenges of ensuring global access to vaccines as the most effective means to reach herd immunity, halt epidemic spread, and save countless lives [51], [52]. While technological innovations have allowed the production of effective vaccines against SARS-CoV-2 in record time, the strategies for delivering these vaccines will be the same that have been implemented around the world for decades: by healthcare professionals, either at health facilities or through outreach activities in the form of vaccination campaigns. Understanding how these delivery strategies can be improved in order to achieve vaccination coverage targets is essential, especially in rural areas of the developing world where delivery is significantly more challenging due to the fragility of health systems and associated health budgets. Using a comprehensive dataset on childhood immunizations at both the health system and population levels in a rural district of Madagascar, we show here how strengthening local health systems can help improve key indicators of vaccination coverage, with different impacts on routine and outreach immunizations. The HSS intervention led to an increase in routine immunizations, resulting in higher vaccination coverage, a reduction in economic inequalities, and a higher proportion of timely vaccinations. Yet, these gains disproportionately benefited those who could most easily access services. Lower contribution of outreach activities in the HSS catchment was associated with a persistence of inequalities in geographic coverage in the area, which prevented achieving international coverage targets for many population groups.

There is widespread agreement that routine immunization should be the basis and the foundation of immunization programmes, but questions remain on how to optimize the delicate balance between providing long-term support to routine immunization and improving short-term access via outreach activities [15] [53]. The multiplicity of barriers to accessing health facilities for populations in low-resource settings, requires mass vaccination campaigns and other outreach activities to maintain or increase coverage, but these strategies can have, in turn, negative effects on the rates of routine immunization [27], [28]. For instance, routine immunization in Madagascar decreased in the months after SIAs and vaccination weeks, resulting in seasonal gaps in immunization and delays from the recommended age of vaccination [27]. Here, we provide complementary insights: where routine immunization improved due to ongoing HSS efforts, the contribution of outreach activities to overall vaccination coverage diminished, with mixed impacts on coverage inequalities. Timeliness of vaccination was better in the HSS catchment, with twice the proportion being vaccinated at the recommended age for polio, DTP and measles in the HSS catchment than in the rest of the district. Timely vaccination is key to ensuring that children are fully protected against common childhood illnesses by the time when they are most at risk, and can help prevent episodic outbreaks [25].

We found that the HSS intervention was associated with a 20-50% increase in the odds of monthly per capita immunizations, which resulted in a 20-40% increase in the odds of coverage per year from 2014 to 2018, and a reduction in economic inequalities over time. This effect may seem counterintuitive, as immunizations are provided free of charge at health facilities across Madagascar as part of the national EPI. However, it has been widely reported that despite childhood vaccines being free of charge, children of poorer households frequently have lower vaccination coverage than their peers [54]–[57], which is consistent with our findings. Seeking health care for healthy children may not be always be a priority for people living under severe poverty, especially given the disproportionate impact of the loss of income associated with seeking care, indirect transportation costs, and lower reported awareness of the long-term benefits of vaccination [54]–[57]. This may explain why BCG vaccination coverage decreased significantly as a function of distance to the health center, as most deliveries in remote areas occur at home. The HSS intervention included, among others, renovations to health facilities, hiring of additional health staff, community sensitization and expanded support for reproductive health, including deliveries in health facilities, antenatal and postnatal care, all of which could have improved the confidence on the health system and increased awareness, particularly among mothers of young children. In addition, the removal of user fees at health facilities, which resulted in a tripling of primary care utilization for individuals of all ages over this period [58], could have had the indirect benefit of increasing health seeking for services that were already free of charge, as adults and mothers get more used to visiting health centers. Health system approaches such as the one implemented in Ifanadiana are increasingly recognized as potential solutions to achieve, not only vaccination coverage targets, but also progress towards universal health coverage [59], [60].

Despite HSS efforts at supporting vaccination delivery at the community level during vaccination weeks and other outreach activities, geographic inequalities in vaccination coverage persisted or even increased for certain vaccines in the HSS catchment, probably as a consequence of the higher contribution of facility-based immunizations. Distance to health care facilities is a known determinant of low vaccination coverage [54], [61], [62], especially in countries like Madagascar, where coverage is lower than average [16]. Outreach activities during vaccination weeks and mass vaccination campaigns can be effective ways to eliminate geographic inequalities [20], as shown in the area of Ifanadiana not supported by the HSS intervention. However, previous modeling studies have shown that eliminating geographic inequalities alone will not achieve coverage targets across Africa, and that parallel increases in routine vaccination rates are necessary [16]. This is consistent with our results, where only certain population groups in the HSS catchment (those of higher socioeconomic level and living in proximity to health centers), but none in the rest of the district, actually reached international coverage targets required for herd immunity. Additional efforts are therefore necessary to sustain improvements in the district, including the geographic expansion of HSS efforts, and a particular focus on outreach activities in the HSS catchment (e.g. more frequent vaccination campaigns, routine expeditions by mobile teams, etc.).

Our study had several limitations. First, we used official MoPH data on population size for children aged 12-23 months in our analysis of per capita immunizations at health centers. These are notoriously inaccurate and can lead to estimated coverage rates above 100%, which was the case in our setting if we had used annualized rates. This is unlikely to have affected our analysis unless inaccuracies in population data were highly structured across health centers (much overestimated in some and underestimated in others). The consistency between health system and cohort results suggests that there was limited bias in the analyses of per capita immunizations. Second, less than one third of the children studied in the cohort had a vaccination card at the time of the interview, so their vaccination status (and therefore estimates of coverage) depended largely on the mother’s report. Although potentially flawed due to recall bias, vaccination coverage figures used by most international organizations and national governments are based on surveys (DHS, MICS, etc.) that use the same methods, and the proportion of children with vaccination cards was not lower here than in other settings [63]. Finally, although the IHOPE cohort includes over 8000 individuals, the sample size for children aged 12-23 months is relatively low, which precludes the robust estimation of vaccination coverage predictors with complex statistical models. For this reason, we expanded the age range of the cohort statistical analyses to children aged 12-59 months. This could have had an impact in the interpretation of results if trends observed for children 12-59 months were greatly different from those in children 12-23.

In conclusion, our study shows that strengthening local health systems can help improve vaccination coverage and timeliness of immunization in rural, low resource settings, even when those interventions do not target specifically vaccine improvements themselves. By increasing the contribution of routine immunization over other immunization strategies such as vaccination weeks or mass campaigns, the intervention helped reduce economic inequalities in vaccination coverage, but failed to reduce geographic inequalities. Overall, the target of 90% immunization coverage was not achieved for any vaccine, but many populations in the HSS intervention area achieved immunization levels above 80%. Explicit efforts are necessary in areas undergoing HSS to vaccinate children in remote areas so that immunization goals can be reached.

## Data Availability

Data used in this study are available from Authors, upon reasonable request

## Table of acronyms

BCG: Bacillus Calmette-Guérin vaccine
DHS: Demographic and Health Surveys
DTP: Diphteria-Tetanus- Pertussis vaccine
EPI: Expanded Program on Immunization
GAVI: Global Alliance for Vaccines and Immunisation
HSS: Health Strengthening System
IHOPE: Ifanadiana Health Outcomes and Prosperity Evaluation
INSTAT: National Institute of Statistics
LMICs: Low and Middle Income Countries
MoPH: Ministry of Public Health
OR: Odds Ratio
RI: Routine Immunization
SIA: Supplementary Immunization Activities
VW: Vaccination weeks

## ACKNOWLEDGEMENTS

The authors acknowledge the contributions and support Herrnstein Family Foundation, and Benjamin Andriamihaja. They are grateful to all of the staff at PIVOT for their field support and their remarkable work in strengthening the health system in Ifanadiana. Thanks are due to the Madagascar Ministry of Health, at both the district and the central levels, for their continuous support and valuable insights. The authors also thank the Institut National de la Statistique (INSTAT) field teams for their involvement in the district wide population survey. Special thanks to Amy Wesolowski for insightful discussions.

## AUTHOR CONTRIBUTIONS

Conceived and designed the experiments: ER, AG, ACM, MHB, CJEM. Collected the data: BRa, MR. Performed the analysis: ER, AG. Wrote the initial draft of the manuscript: ER, AG. Revised the manuscript and accepted it in its final form: ER, MHB, ACM, FI, LFC, BRaz, FHR, KEF, RJLR, GC, Brat, FR, MR, EMRF, AR, CJEM, BRo, AG.

## SUPPLEMENTARY INFORMATION

**S1 Supplementary information appendix**. It contains 2 supplementary tables and 1 figure.

