## Appendix for "Impact of health system strengthening on delivery strategies to improve child immunization coverage and inequalities in rural Madagascar"

- Supplementary information Appendix -

Elinanbinina Raojaonarifara^1,2,3^, Matthew H. Bonds^2,4^, Ann C Miller^4^, Felana Ihantamalala ^2,4^, Laura F Cordier^2^, Benedicte Razafinjato^2^, Feno H. Rafenoarimalala^2^, Karen E. Finnegan^2,4^, Rado J. L. Rakotonanahary^2^, Giovanna Cowley^2^, Baolova Ratsimbazafy^2^, Florent Razafimamonjy^2^, Marius Randriamanambintsoa^5^, Estelle M. Raza-Fanomezanjanahary^6^, Andriamihaja Randrianambinina^6^, C. Jessica E. Metcalf^7^, Benjamin Roche^1^, Andres Garchitorena^1,2^

*^1^ MIVEGEC, Univ. Montpell,ier, CNRS, IRD, Montpellier, France*

*^2^ NGO PIVOT, Ranomafana, Madagascar*

*^3^ Department of Global Health and Social Medicine, Harvard Medical School, Boston, USA*

*^4^ Direction de la Démographie et des Statistiques Sociales, Institut National de la Statistique, Antananarivo, Madagascar*

*^5^ Ministry of Public Health, Antananarivo, Madagascar*

*^6^ Sorbonne Université*

*^7^ Department of Ecology and Evolutionary Biology, Princeton University, Princeton, NJ, USA*

*^8^ Department of International Health, Johns Hopkins University, Baltimore, Maryland, USA*

**Table S1.** Summary of the idHSS intervention carried out by the MoPH-PIVOT partnership in Ifanadiana District in 2014-2017, based upon TIDieR guidance

**1. BRIEF NAME**

Integrated district-level health system strengthening (idHSS) initiative in Ifanadiana, Madagascar

**2. WHY**

***GOAL/RATIONALE:*** To create a model public health district with universal access to care aiming for broad-based population health impact on mortality. Based upon the World Health Organization’s building blocks of Health System Strengthening: 1) service delivery; 2) health workforce; 3) health information systems; 4) medicines and supplies; 5) financing; 6) leadership.

**WHAT**

**3. *MATERIALS*** (by level of care and above enumerated building blocks)

**At district hospital level**

(1) *Service delivery:* Overall infrastructure upgrades and outfitting for service delivery, bringing inpatient bed capacity from 25 to 40, upgrades to waste management system, specific renovations of the emergency and triage department, pediatric unit, inpatient ward, isolation ward, and laboratory; support to specific service delivery, including emergency care and provision of a network of 3 ambulances, including 2 new fully equipped ones with 24/7 coverage and 12 PIVOT paramedics for referrals; maternal and obstetrical care; laboratory service were upgraded to include a total of 53 tests, including microscopy and GeneXpert for tuberculosis; social support evolved to support all hospitalized and vulnerable patients; launch of intensive care unit for severe acute malnutrition with complications.

(2) *Health workforce:* Staffing of health workers to reach MoPH norms through joint MoPH-PIVOT hires of 7 clinicians, including a trauma surgeon and an anesthesiologist, which were integrated into the MoPH staff (long term solution); staffing supplemented further with fulltime presence of 2 PIVOT doctors and 4 nurses by end of 2017; non-clinical PIVOT staff including a team of 3 social workers, support staff (janitors, guards, etc.), a laboratory technician, and a radiology technician; ongoing mentorship and frequent trainings of medical staff in key clinical areas, such as emergency medicine and postoperative care.

(3) *Health information systems:* Creation of a hospital-based M&E team to follow progress of activities and improve quality of HMIS data; implementation of a system for baseline and follow-up facility readiness surveys.

(4) *Medicines and supplies:* Supply chain management and reduction of stock-outs, initially through frequent donations which evolved into a reimbursement program paired with pharmacy management training; provision of medical and non-medical equipment for service delivery, including full laboratory capacity. PIVOT became the procurement manager for the hospital pharmacy as of October 2017.

(5) *Financing:* Cost of outpatient and inpatient care fully covered for patients referred by district-wide health centers and self-referred patients who necessitated urgent inpatient care (over 76,000 patients between 2014 and end of 2017); cost of referral to and care at higher levels of care (e.g. university hospital) fully covered for services not available at district hospital.

(6) *Leadership:* Creation of a joint MoPH-PIVOT executive committee for hospital management and transparency; creation of sub committees for specific projects such as infection control or quality of care.

**At health center level**

(1) *Service delivery:* Overall infrastructure renovations and/or extensions for service delivery at 5 target health centers, including ensuring electricity, water, waste management/sterilization capacity, proper pharmacy conditions; provision of medical and non-medical equipment, including beds, armoires, furniture; support to launching specific service delivery of Integrated Management of Childhood Illnesses (IMCI) and malnutrition protocols for every child under 5 attending the health center; ensuring timely referrals and emergency care. Launch of supervision efforts and quality of care improvement projects with a focus on IMCI and malnutrition.

(2) *Health workforce:* Staffing through joint MoPH-PIVOT hires to bring all 13 primary care health centers up to MoPH norms (1 doctor, 1 nurse, 1 midwife, 1 dispenser, 1 support staff at each facility); 33 PIVOT-MOH hires were integrated into the Ministry of Health staff (long term solution); at target health facilities, hiring exceeded norms and PIVOT clinicians were permanently present (~2 clinicians per health center) to implement service delivery protocols (e.g. IMCI, malnutrition); trainings for medical staff (some district-wide) such as obstetrical and neonatal care; ongoing supervision and mentorship in target centers for IMCI and malnutrition.

(3) *Health information systems:* Joint MoPH-PIVOT training and supervision to improve HMIS data quality (district-wide); implementation of system for baseline and follow-up of facility readiness surveys.

(4) *Medicines and supplies:* Supply chain management and reduction of stock-outs, initially through frequent donations which evolved into a reimbursement program paired with pharmacy management training.

(5) *Financing:* Essential medicines and consumables provided free of charge to all patients (more than 130,000 patients between October 2014 launch and end of 2017); more details of this program are available in Section S1 of the Appendix.

(6) *Leadership:* Close collaboration with district health managers for the planning and implementation of activities.

**At community level**

(1) *Service delivery:* Construction of 21 community health posts; specific service delivery in Integrated Management of Childhood Illnesses and malnutrition protocols for every child under 5, community sensitization and mass testing, urgent care, and mobile clinics with direct care provision by PIVOT clinicians every other month. In 2017 PIVOT started to support monthly supervision of CHWs at the health facilities and doubled the area of the CHW strengthening program (43 community health posts, covering 4 out of 5 communes in the idHSS catchment).

(2) *Health workforce:* 14 active CHW supervisors – PIVOT staff that are training, coaching and monthly supervision of ~86 community health workers by mobile teams of trained nurses by the end of 2017; community IMCI training provided for CHWs in all of the intervention area

(3) *Health information systems:* Joint MoPH-PIVOT training to improve HMIS data quality on community health.

(4) *Medicines and supplies:* Monthly provision and follow-up of MNCH medicine stocks, including malaria diagnosis and treatment, oral rehydration salts, NSAIDS, antibiotics and zinc.

(5) *Financing:* cost of MNCH medicine stocks fully covered; financial and non-financial incentives to CHWs and local leadership.

(6) *Leadership:* Community engagement and participation (e.g. community health posts are built by the community, with PIVOT support for roofing, painting, furniture and equipment).

**4. *PROCEDURES***

All interventions were aimed at fulfilling existing Madagascar Ministry of Health protocols and standards.

**5. WHO PROVIDED**

**At district hospital level**

Ministry of Health clinicians provided the majority of service delivery. PIVOT clinicians are integrated in the hospital staff and provide direct care as any other clinician during external consultations and clinical rounds, but also carry out frequent training. Non-clinical PIVOT staff provided social support to vulnerable patients, helped manage the patient circuit to benefit from fee exemptions (registration, validation).

**At health center level**

Ministry of Health doctors and nurses provided the majority of service delivery. By MoPH norms each health center (CSB2) should have 1 doctor, 1 nurse, 1 midwife, 1 dispenser, and 1 support staff. Additional PIVOT clinicians (~2 per health facility) provided some direct care, especially for the implementation of malnutrition and IMCI protocols, but focused mostly on training and supervision.

**At community level**

Two community health workers per fokontany (a cluster of villages, lowest administrative unit) provided basic MNCH care, supervised monthly by the clinicians of their respective health center. PIVOT mobile teams of nurses provided on-site mentoring and supervision of CHWs every two months. They also provided direct care at community level during on-site supervisions for fokontany located >10 kilometers from a health center.

**6. HOW (modes of delivery)**

PIVOT employees worked in partnership with existing networks of MoPH clinicians and community health workers within existing public health facilities. Wherever possible, such as in the case of supply chain management, leadership and financing, the intervention deliberately avoids the creation of parallel systems of care.

**7. WHERE**

**At district hospital level**

The initial PIVOT catchment area comprised the only district hospital, located in Ifanadiana city. Most referrals to higher levels of care (tertiary) were sent to the university hospital in Fianarantsoa (2h away by car), and some to specialized facilities in Antananarivo (capital, ~1 day by car).

**At health center level**

Full health center activities were implemented in the health centers of the five communes closest to the hospital on the district’s sole paved road (i.e. Ranomafana, Kelilalina, Ifanadiana, Tsaratanana, Antaretra); all 13 health centers in the district received trainings, staffing support to reach MoPH norms, and some access to the referral network (limited by road conditions and accessibility).

**At community level**

By the end of 2017, community activities had been rolled out in fokontany from four of the five communes within the PIVOT catchment area (43 out of a total of 195 total fokontany in the district).

**8. WHEN AND HOW MUCH**

All interventions were progressively rolled out during the study period.

The earliest intervention activities implemented (starting in April-May 2014, at the beginning of the study period) included the ambulance network, staffing of health centers and district hospital, and provision of medical equipment in four communes.

The renovation of health centers also began in April-May 2014 but the date of completion varied for each health facility.

The renovation of the emergency and triage unit and pediatric guard at the district hospital were completed by early 2016.

Removal of user fees at health centers and hospital began in October of 2014.

Implementation of IMCI and malnutrition protocols at all health centers began in October 2015.

Community-level activities began in November 2015 in two communes, with an expansion to four communes in February 2017.

First expansion of the PIVOT intervention area at health facility level to include a fifth commune in October 2017.

A costing analysis of the idHSS intervention is underway and will be published separately. Preliminary estimates suggest that the annual per capita investment of the idHSS intervention (all in-country costs of the NGO/idHSS catchment population) was about $35, which added to the investment by the MoPH and other bilateral or multilateral donors represents approximately $65 per capita.

**9. TAILORING**

The idHSS intervention in Ifanadiana District was initially modelled after the experience in HSS implementation by the NGO Partners in Health in several districts of Rwanda. A distinct element of the idHSS intervention was a substantial investment in information systems, monitoring, evaluation and research as core elements of the intervention. For M&E, an interactive dashboard allows the visualisation of hundreds of indicators from health system data collected at all levels of care. Key indicators are reviewed in monthly and quarterly reviews with program managers to follow-up the progress of different activities and services (e.g. utilization, quality of care). For research, the I-HOPE cohort study allows conducting impact evaluations every two years, which provides complementary population-level information such as health system coverage, quality, or mortality. Evaluation results are presented to managers and leadership of the MoPH-PIVOT partnership as they become available, but to allow the routine use of this information by program managers, an interactive web application has been developed (main text).

As a result of the iterative learning process integrated in the idHSS intervention, program implementation has been tailored over time to respond to coverage gaps and intervention deficiencies observed in Ifanadiana during monitoring, evaluation and research activities. The most notable example of this is the increasing support to community health after an initial phase where the idHSS intervention focused most of its resources on strengthening the district hospital and several health centers. The data and analyses generated by the I-HOPE cohort study (among other sources) have contributed to this shift, highlighting geographic inequalities in coverage and leading to several programs to address them. First, PIVOT has expanded its support to the community health program, both geographically and in terms of activities implemented. In 2019-2020, PIVOT is piloting the implementation of proactive community health in order to improve access to care for children under 5 years to further reduce geographic barriers in access to care within the communities. Second, the NGO and local government are building houses near health centers for mothers to arrive several days in advance of their delivery date in order to increase geographic access to safe delivery. This is part of a broader effort to improve maternal health coverage, given lags in maternal care improvements observed in the idHSS catchment. Third, PIVOT is increasing its outreach activities, such as on-site supervision of community health workers by teams of nurses (who also provide direct care during field expeditions) and is looking into progressively expanding the scope of work and professionalization of community health workers.

**10. MODIFICATIONS**

N/A; The intervention is progressively being implemented, as explained in section 8 (when and how much)

**HOW WELL**

N/A; The aim of this study was to study the evolution of geographic access to primary care in Ifanadiana District. Full details of the impact assessment are available in the main text.

**Table S2.** Vaccination coverage in Ifanadiana for each of the recommended vaccines, 2014-2018

| Variables | Statistics of the coverage vaccination (%) | | | | | | | | | | | | | | |
| --- | --- | --- | --- | --- | --- | --- | --- | --- | --- | --- | --- | --- | --- | --- | --- |
|  | 2014 | | | | | 2016 | | | | | 2018 | | | | |
|  | BCG | Measles | DTP | Polio | All | BCG | Measles | DTP | Polio | All | BCG | Measles | DTP | Polio | All |
| All      **Socio-economic class**  - Poorest (Q5)  - Second poorest (Q4)  - Middle (Q3)  - Second wealthiest (Q2)  - Wealthiest (Q1)  **Distance to health center**  - <5km  - 5km<<10km  - >10km  I **HSS catchment**  - outside  - inside | 53.8  51.7  53.7  58.6  44.2  61.9  66.8  43.3  40.0  55.3  50.9 | 57.4  43.2  57.8  56.6  52.1  81.1  71.4  50.1  26.8  54.6  62.8 | 57.5  53.3  51.4  46.4  58.9  80.3  72.3  50.6  21.2  51.9  67.9 | 59.2  56.6  49.0  53.5  57.7  83.1  75.9  51.5  18.0  55.2  66.7 | 34.6  30.0  28.9  32.0  29.0  56.2  52.9  22.3  05.1  37.6  32.9 | 68.2  75.8  66.2  54.9  69.4  81.2  71.3  65.3  64.1  64.7.8  73.9 | 61.7  75.0  55.4  43.9  59.8  85.9    70.7  52.0  53.2    54.8  73.1 | 60.3  56.7  53.0  52.0  62.3  87.9  70.2  51.7  45.5  53.3  71.7 | 70.7  75.4  60.2  67.2  68.2  90.2    78.6  63.5  59.8  66.3  78.1 | 41.3  44.7  37.8  24.6  44.8  65.0  49.3  33.2  32.1  35.8  50.4 | 74.3  70.5  71.7  60.5  91.1  87.0  78.2  68.6  73.9  70.3  80.8 | 58.3  42.3  51.4  53.5  77.1  75.5  63.3  53.0  52.1  49.2  73.2 | 69.5  61.5  60.8  65.1  85.3  81.2  72.1  68.0  62.4  65.7  75.6 | 74.2  71.5  67.6  68.1  91.3  78.3  79.5  70 .5  62.0  71.3  79.0 | 47.4  34.0  36.6  40.6  68.7  66.3  59.73  42.62  39.70  37.5  63.6 |

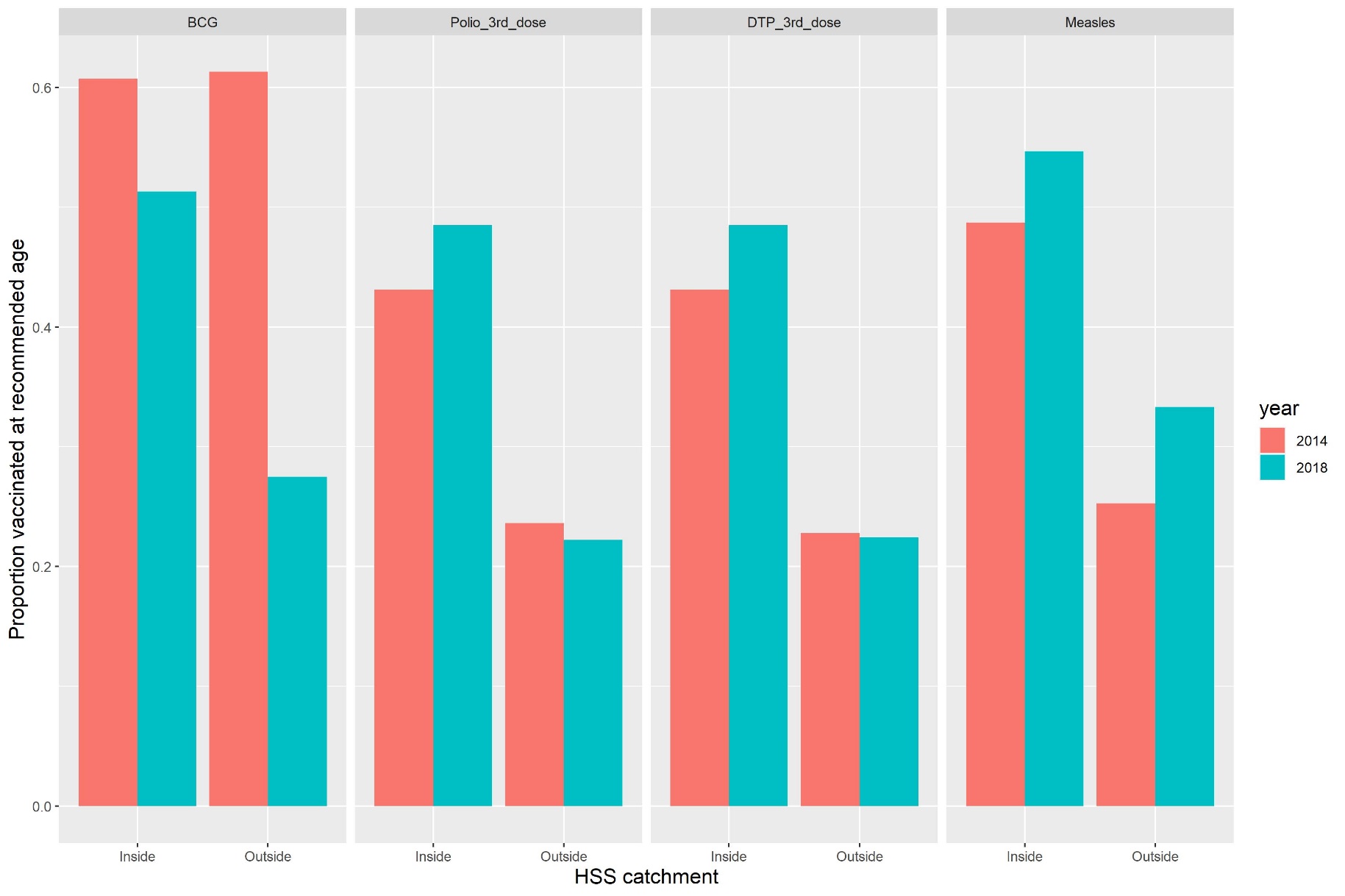
**Figure S1.** Timeliness of vaccination over time for each of the recommended vaccines in Ifanadiana District, inside and outside the HSS catchment.
